# The impact of EEG preprocessing parameters on ultra-low-power seizure detection

**DOI:** 10.1101/2025.01.17.25320722

**Authors:** Patrick Reisinger, Jonathan Larochelle, Ciamak Abkai, Sotirios Kalousios, Nico Zabler, Matthias Dümpelmann, Andreas Schulze-Bonhage, Peter Woias, Laura Comella

## Abstract

**Objective:** Closed-loop neurostimulation is a promising treatment for drug-resistant focal epilepsy. A major challenge is fast and reliable seizure detection via electroencephalography (EEG). While many approaches have been published, they often lack statistical power and practical utility. The use of various EEG preprocessing parameters and performance metrics hampers comparability. Additionally, the critical issue of energy consumption for an application in medical devices is rarely considered. Addressing these points, we present a systematic analysis on the impact of EEG preprocessing parameters on seizure detection performance and energy consumption, using one to four EEG channels.

**Methods:** We analyzed in 145 patients with focal epilepsy the impact of different sampling rates, window sizes, digital resolutions and number of EEG channels on seizure detection performance and energy consumption. Focusing on clinically relevant, event-based metrics, we evaluated seizure detection performance of a state-of-the-art convolutional neural network (CNN) via the Seizure Community Open-Source Research Evaluation (SzCORE) framework. Statistical relevance of parameter changes was assessed using linear mixed-effects models. Energy consumption was analyzed using an ultra-low-power microcontroller.

**Results:** Reducing the sampling rate from 256 to 64 Hz led to a decrease in sensitivity (*p* = .015) and false detections per hour (FD/h; *p* = .002). Larger window sizes reduced sensitivity between 1 and 8 second windows (*p* = .033) and FD/h between one second and all other sizes (all *p* values < .001). Average detection delays increased between one second and 4 and 8 second windows (both *p* < .01). Lower digital resolutions decreased sensitivity between 16 and 8 bits (*p* = .007). Compared to four channels, using only one EEG channel resulted in a decrease in sensitivity (*p* < .001) and an increase in the average detection delay (*p* = .020), but showed less FD/h (*p* = .005). CNN energy consumption decreased from 49.15 to 17.26 µJ/s when the sampling rate was reduced from 256 to 64 Hz. Lowering the number of channels from four to one reduced the CNN energy consumption from 79.04 to 31.63 µJ/s.

**Significance:** This study provides guidance on choosing EEG preprocessing parameters for innovative developments of closed-loop neurostimulation devices to further advance the treatment of drug-resistant focal epilepsy.

**Key points:**

- A major challenge in closed-loop neurostimulation is fast and reliable seizure detection, ideally with minimal energy consumption.
- Many published seizure detection approaches are not ready for application and lack statistical power and comparability.
- The impact of EEG preprocessing parameters was systematically evaluated in 145 patients with focal epilepsy.
- The chosen window size and number of channels had the strongest impact on seizure detection performance and energy consumption.
- Our results offer guidance for EEG preprocessing choices to advance the treatment of drug-resistant focal epilepsy.

## Introduction

In children and adults with epilepsy, focal epilepsy is the most common form (1), which places a great burden on those affected, caregivers and healthcare systems (2). Although there are several antiseizure medications on the market, one third of individuals with epilepsy show drug-resistance (3,4), with higher prevalence in focal epilepsy (5). An alternative therapy option for drug-resistant focal epilepsy is neurostimulation, where invasive approaches such as vagus nerve stimulation (VNS), deep brain stimulation (DBS) and responsive neurostimulation (RNS) exist (6,7). A novel, minimally invasive approach is the EASEE® system (Precisis GmbH, Heidelberg, Germany), where a thin electrode is placed subcutaneously over the epileptic focus (8,9). In order to improve therapeutic efficacy, closed-loop capabilities are increasingly considered in neurostimulation devices (10).

One major challenge in closed-loop neurostimulation is fast and reliable seizure detection, in order to trigger stimulation pulses, which could prevent the further spread or even the onset of seizures (11,12). Over the last decades, seizure detection based on electroencephalography (EEG) data with annotated seizures has been extensively studied (13–16). The most common approaches are deep learning techniques such as convolutional (17) or recurrent neural networks (18), as well as machine learning algorithms such as random forest (19). These algorithms are usually trained and evaluated with a patient-specific approach, because of its higher performance compared to a patient-generic approach (20).

However, many of these approaches are not ready to use in a neurostimulation device. First, only a small number of EEG channels can be used for detection, since constantly wearing an EEG cap is impractical and stigmatizes the patient (21). Although there are many minimally visible EEG devices on the market (22), a fully implanted device is often more suitable for closed-loop neurostimulation, such as DBS where subcortical brain regions are targeted (10). Next, the power consumption of seizure detection models is usually not investigated, although battery lifetime is one of the central factors enabling seizure detection on ultra-low-power devices (20). Extending the battery lifetime decreases the frequency of surgical procedures required for device replacement, thereby reducing both healthcare costs and patient risk (23). Furthermore, most seizure detection models are evaluated using the openly available CHB-MIT Scalp EEG Database, consisting of EEG recordings from a small group of 22 pediatric patients (24,25). Since epileptic seizures have different EEG characteristics in children (26), generalizability of results obtained with this database to adult patients is limited. Many publications use conventional sample-based evaluation metrics, which tend to show high performance in imbalanced datasets (27). While these studies provide valuable insights, it is advisable to use evaluation metrics that incorporate enhanced clinical and practical relevance. For real-world applications, an event-based approach such as the recently proposed Seizure Community Open-Source Research Evaluation framework (SzCORE) is therefore more suitable (28). Furthermore, appropriate statistical testing is seldom applied when comparing different models (29) and statistical power is in general reduced when using the small CHB-MIT database. In addition to that, many variations of EEG preprocessing parameters are used and only a limited number of studies investigated their impact on seizure detection performance and energy consumption (20,30). Commonly varied parameters are window size (31), sampling rate (32) and number of channels (33). To our best of knowledge, no work exists that evaluates the effects of varying the digital resolution of EEG data.

Here, we systematically analyze the impact of different EEG preprocessing parameters on patient-specific seizure detection performance and energy consumption in a large group of 145 patients with focal epilepsy (Figure 1). We deployed a state-of-the-art convolutional neural network on an ultra-low-power microcontroller and statistically tested the impact of different sampling rates, window sizes, digital resolutions and number of channels using linear mixed-effects models. We show that ultra-low-power seizure detection is feasible with low sampling rates, small window sizes, low digital resolutions, as well as using only one EEG channel. All analyzed parameters had their own unique impact on seizure detection performance and energy consumption. The overall goal of this study is to provide guidance on the choice of EEG preprocessing parameters for the development of closed-loop neurostimulation to further advance the treatment of drug-resistant focal epilepsy.

**Figure 1.**
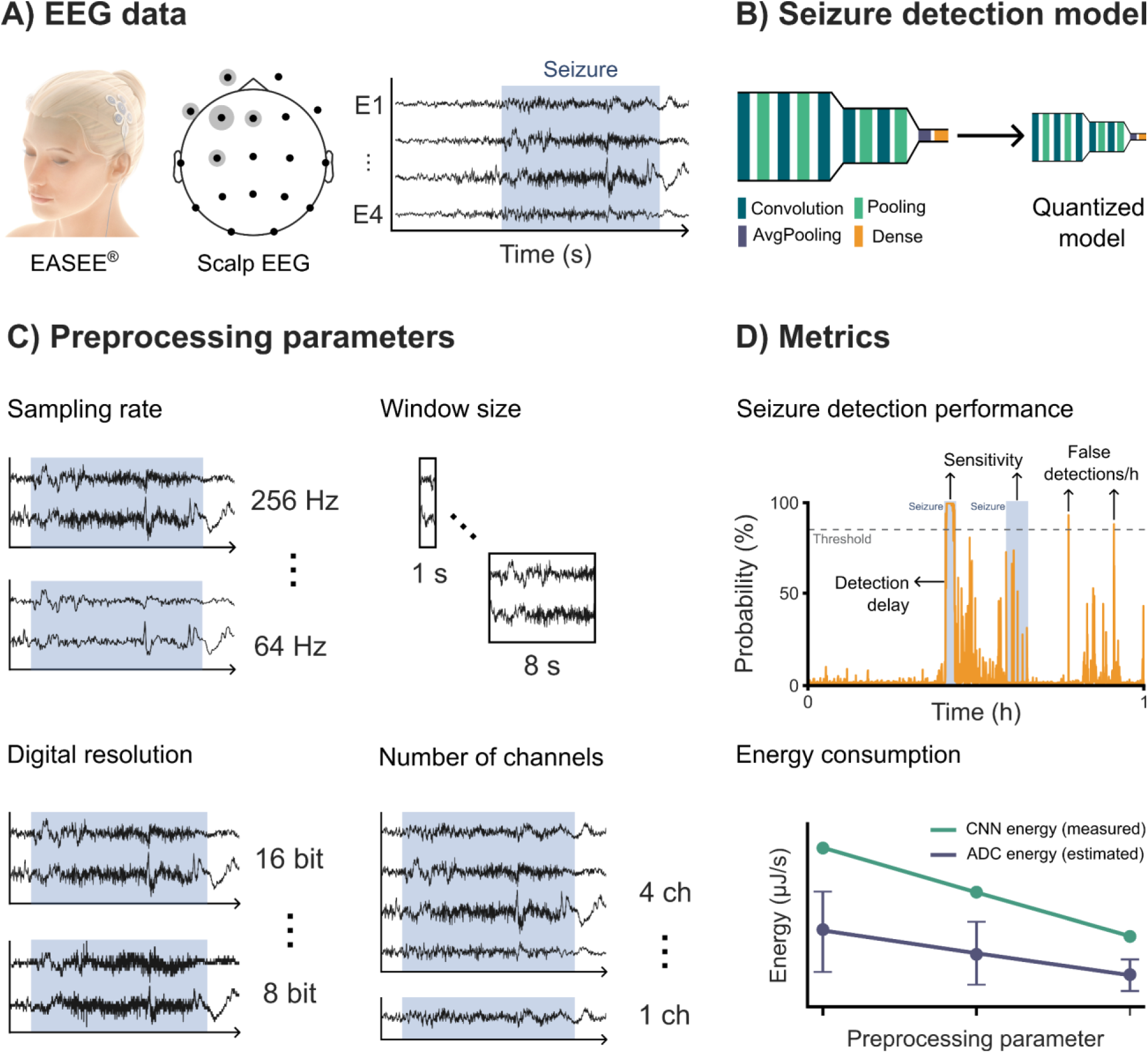
Overview of study and analysis rationale. (A) To mimic the electrode of the EASEE® system, scalp EEG data is reduced to a central channel at the epileptic focus and its four closest channels. Example EEG data with an annotated seizure is shown for four channels re-referenced to the central channel. (B) A convolutional neural network (CNN) is used for seizure detection, which is quantized in order to be executed on a microcontroller. (C) Examples of the different used sampling rates, window sizes, digital resolutions and numbers of channels from the highest to the lowest value. (D) Seizure detection performance was evaluated using event-based scoring, with a focus on sensitivity, detection delay and false detections per hour. CNN energy consumption was measured across each varied parameter and analog to digital converter (ADC) energy was estimated.

## Materials & Methods

The following is an abbreviated description. Further details for each section can be found in the supplementary material.

### EEG database and preprocessing

We used surface EEG recordings of the EPILEPSIAE database (34) from a total of 150 patients with focal epilepsy (35). General EEG preprocessing was done using MNE-Python (36) and custom scripts. When necessary, the EEG data was downsampled to 256 Hz. To remove the DC offset and power line noise, a bandpass filter was applied between 0.5 and 48 Hz. A previous study demonstrated that sub-scalp EEG recorded via the electrode of the EASEE® system is highly similar to surface EEG recordings (37). To mimic the electrode configuration of the EASEE® system with the surface EEG data used in this study (38), the number of electrodes was reduced to one central and four additional peripheral electrodes covering the seizure onset zone (see Figure 1A). Subsequently, the data was re-referenced to the central electrode, resulting in four EEG channels.

### Seizure detection model

Seizure detection was modeled as a binary classification task and trained using TensorFlow and Keras. We adapted *EpiDeNet* (39), a lightweight seizure detection convolutional neural network (CNN). A model using 2-channel EEG with a sampling rate of 256 Hz, a window size of one second (with 0% overlap) and a digital resolution of 16 bits served as reference for comparison (see Table 1). For computational reasons, we only changed the preprocessing parameters in one dimension and compared them with the other parameters investigated in the corresponding dimension.

**Table 1.**
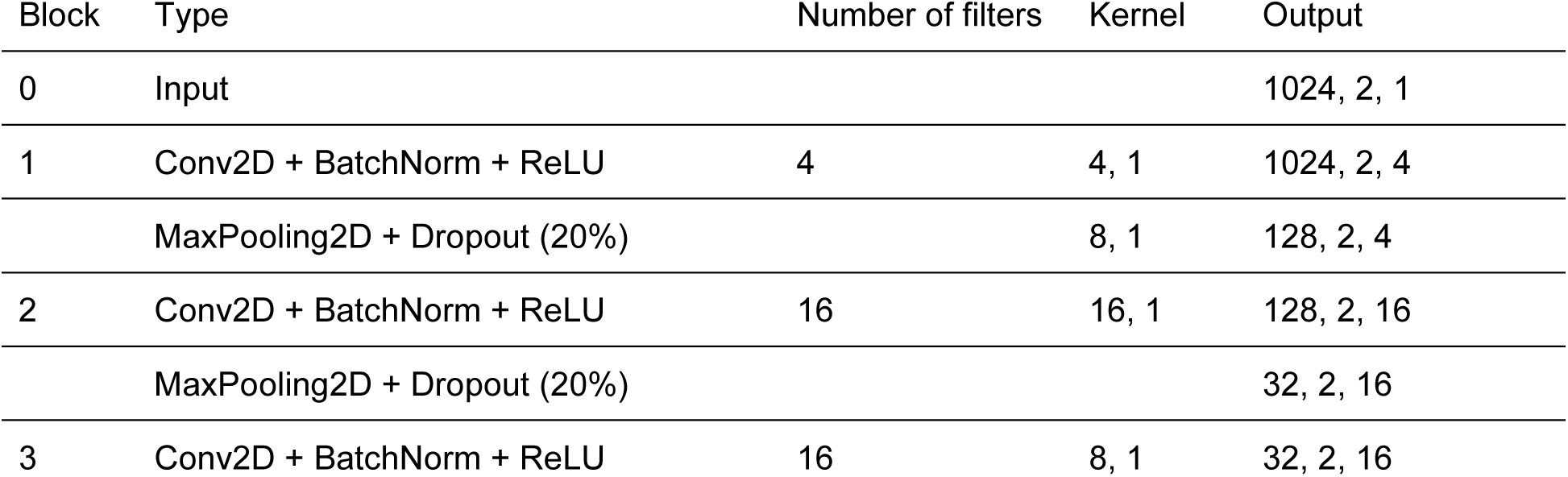
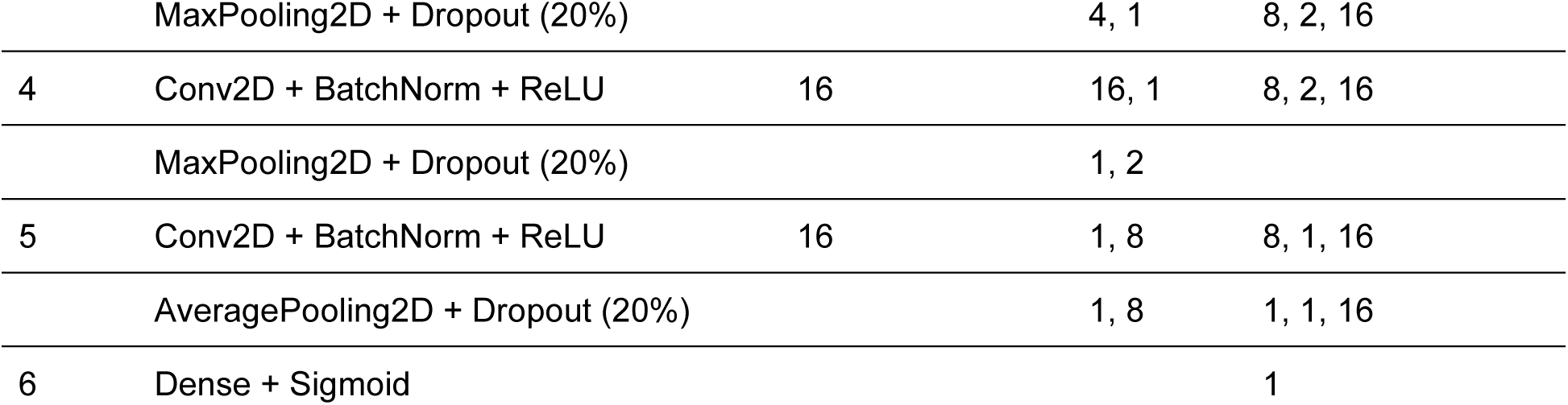
CNN architecture adapted from EpiDeNet (39) for the baseline model of this study.

Since the focus of this study is on patient-specific models, time-series cross-validation is applied to preserve the chronological order of recordings and prevent performance overestimation (28). In each cross-validation step, the next chronological training data segment is added, and the subsequent segment is used as testing data for performance evaluation. This process was repeated until all segments were used per patient, which were on average 5.7 segments. Segments were one hour long and contained at least one seizure. All data was normalized based on a scaler (z-scoring) fitted using the training data. Additionally, the windowed training data and its corresponding labels were shuffled.

### Sampling rate

Sampling rates of 256, 128, and 64 Hz were investigated to determine their impact on seizure detection and energy consumption. CNN parameters for 256 and 128 Hz were changed in the first input dimension, corresponding to the number of samples. When using a sampling rate of 64 Hz, kernel sizes were also adapted.

### Window size

We also investigated different window sizes and their impact on seizure detection performance and energy consumption. In this study, we considered non-overlapping window sizes of 1, 2, 4, and 8 seconds. Additionally, windows with an overlap of 50% were also analyzed. CNN parameters were adapted in the first input dimension, ranging from 256 samples in 1 second to 2048 samples in 8 seconds.

### Digital resolution

The digital resolution, in bits, is given by the EEG analog to digital converter (ADC) which samples the analog raw signal. The data used for this study has a digital resolution of 16 bits. Digital resolutions of 16, 14, 12, 10, and 8 bits were analyzed regarding their effects on seizure detection performance and energy consumption. No adjustments of CNN parameters were conducted, since the input sizes did not change compared to the baseline model.

### Number of channels

Another major preprocessing parameter is the number of used EEG channels. In this study, seizure detection performance and power consumption were evaluated using as few as one channel and up to a maximum of four channels, all re-referenced to a central channel. We used a pseudo-randomized scheme and labeled the initially selected four channels as E1 to E4. These channels represent the closest four channels to the seizure onset zone within each participant. When using three channels, we kept the channels with the labels E2, E3 and E4. When using two channels, we kept E1 and E3. In the case of one channel, we trained and evaluated the models for each patient using E1 to E4 once to minimize bias towards one of the channels. We report the seizure detection performance metrics averaged across these four one-channel models. CNN parameters were changed in the second input dimension, corresponding to the number of EEG channels.

### Seizure detection performance

For evaluating the performance of the seizure detection, we followed the guidelines and recommendations of the recently proposed SzCORE framework (28). In this study, we focus on event-based performance metrics that are more relevant for practical applications compared to traditional sample-based metrics (40). Specifically, we analyzed sensitivity, false detections per hour and average detection delay based on the averaged metrics obtained per patient. Since we used a pre-ictal tolerance of up to 30 seconds, as advised by SzCORE, the average detection delay can also be negative. Furthermore, the classification threshold that maximizes the F1 score of the training data was chosen. All reported metrics are based on models quantized to 8 bits with TensorFlow Lite to enable a more realistic performance evaluation for a neurostimulation application.

### Energy consumption

To characterize the energy consumption, we consider the sum of ADC sample energy and CNN energy. We measure the CNN energy on an EFR32MG24 microcontroller, which is able to accelerate CNNs (Silicon Laboratories Inc., Austin, USA) and report it as mean CNN energy per second. ADC energy cannot be trivially obtained, because different commercially available chips are optimized for different digital resolutions and sampling rates. Therefore, it is estimated using Boris Murmann’s ADC Performance Survey (41) and reported as median ADC energy per second. The corresponding interquartile ranges (IQR) and further details on CNN energy are reported in the supplementary material.

### Statistical analysis

To statistically compare the impact of preprocessing parameters on seizure detection performance while accounting for the structure of the data, we calculated linear mixed-effects models (LMMs) in R using the lmerTest package. The fixed effect for the preprocessing parameter was entered into the model as an ordered factor, with 256 Hz as the reference value for the sampling rate, 1 second for the window length, 16 bits for the digital resolution and 4 for the number channels. For each model, we report regression coefficients (*β*) and p values (*p*), separately for each statistically significant comparison with the reference value of the respective preprocessing parameter. For all statistical tests, a significance level of α = .05 was used. Full model metrics and additional post-hoc contrasts are reported in the supplementary material.

## Results

From the initial 150 patients, five patients were excluded due to either too short seizure EEG data, flat channels, incorrect timestamps or incomplete datasets. This resulted in a final sample size of 145 patients with a total data length of 2229 hours, including 1126 annotated seizures, equivalent to 21 hours of seizure data. The reported seizure detection performance metrics are dependent on the chosen threshold when an event is classified as a detection. Across all investigated parameters, this classification threshold was on average 83.0%, with a standard deviation of 11.5%. Due to substantial interindividual variability in the seizure detection performance metrics, median values are reported in the following sections (for detailed metrics see supplementary tables S12-16).

### Impact of sampling rate

First, we compared the impact of using different sampling rates on sensitivity (Figure 2). Median sensitivities for 256 Hz were 83.3%, 72.7% for 128 Hz and 66.7% for 64 Hz. There was a significant difference from 256 Hz in 128 Hz (*β* = −5.70, *p* = .008) and 64 Hz (*β* = −5.16, *p* = .015).

**Figure 2.**
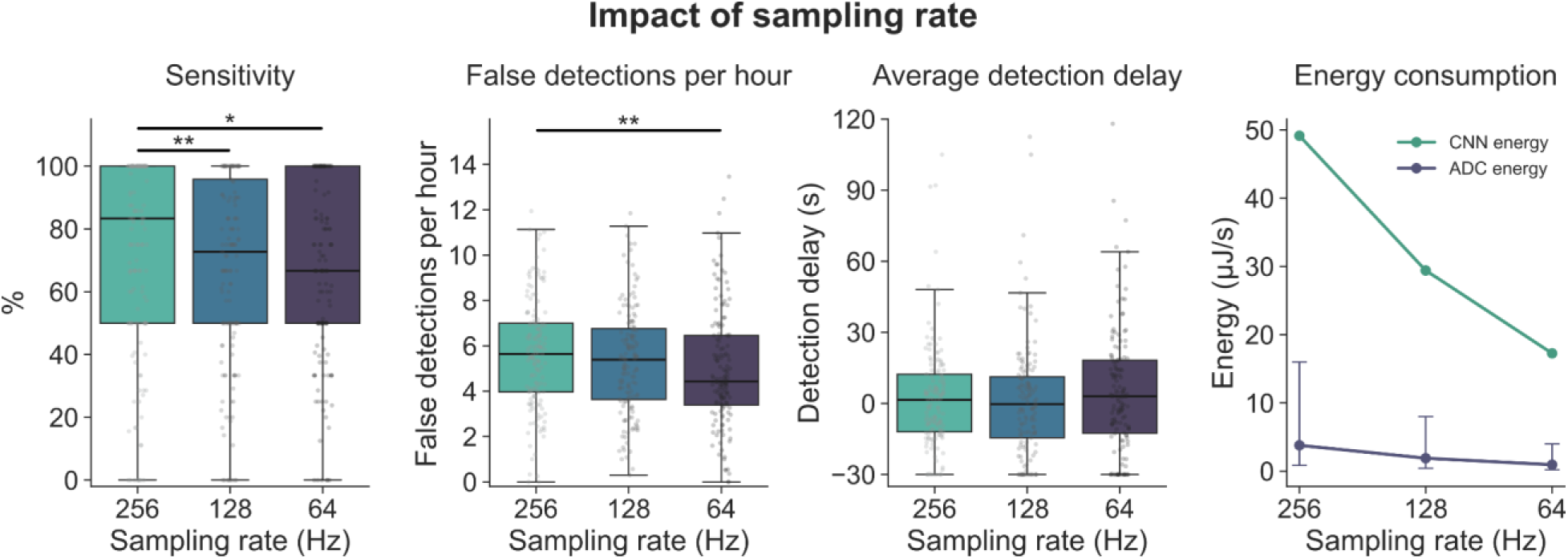
Impact of sampling rate on performance metrics and energy consumption. The gray points represent the individual patients. The error bars for ADC energy consumption represent the IQR. \**p* < .05, \*\**p* < .01

Median false detections per hour were 5.6 FD/h for 256 Hz, 5.4 FD/h for 128 Hz and 4.4 FD/h for 64 Hz, with a significant difference between 256 Hz and 64 Hz (*β* = −0.64, *p* = .002).

The average detection delay showed a median of 1.6 s when a sampling rate of 256 Hz was used. With a sampling rate of 128 Hz, the average detection delay was −0.3 s and 3.0 s for 64 Hz. There was no significant difference.

The variation of the sampling rate showed that the mean CNN energy consumption was 49.15 µJ/s for 256 Hz, 29.41 µJ/s for 128 Hz and 17.26 µJ/s for 64 Hz. The median ADC energy consumption was 3.79 µJ/s when using 256 Hz, 1.89 µJ/s for 128 Hz and 0.95 µJ/s for 64 Hz.

### Impact of window size

Next, we analyzed the impact of the chosen window size (Figure 3). The median sensitivity for 1 second windows was 83.3%, 75.0% for 2 seconds, 80.0% for 4 seconds and 66.7% for 8 seconds. There was a significant difference in sensitivity when comparing the 1 second window size with 8 seconds (*β* = −4.06, *p* = .033).

**Figure 3.**
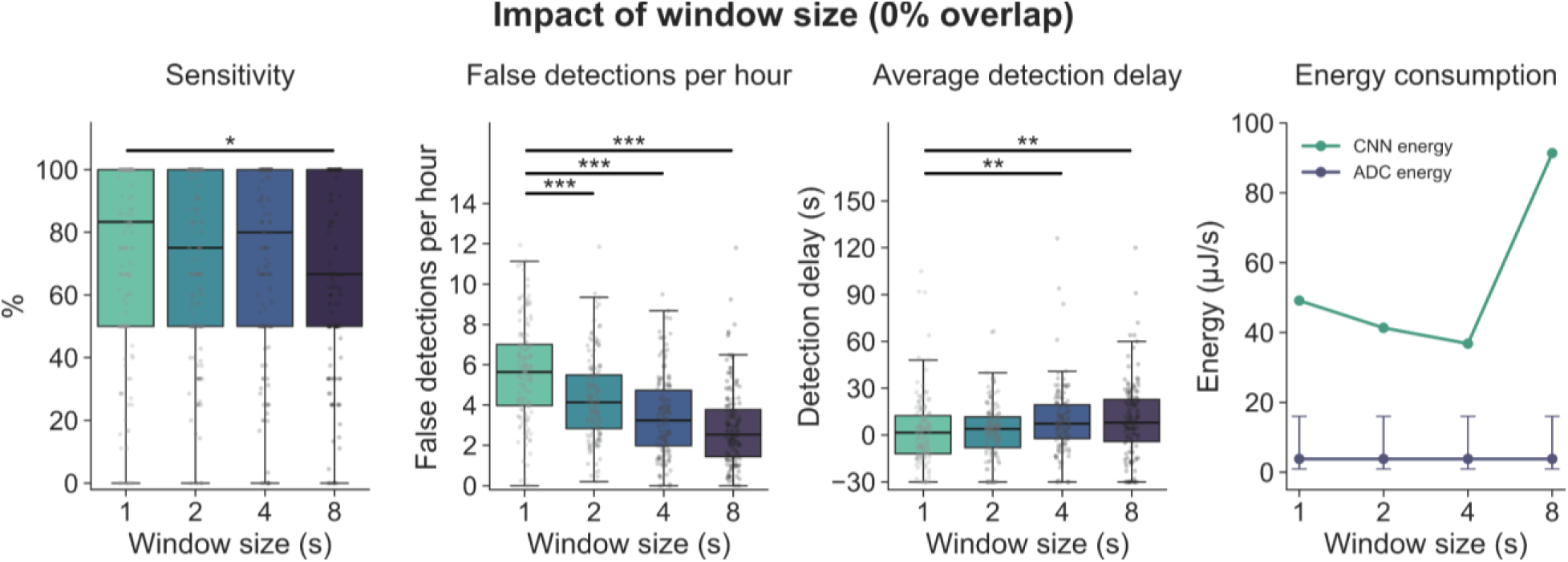
Impact of window size with 0% overlap on performance metrics and energy consumption. The gray points represent the individual patients. The error bars for ADC energy consumption represent the IQR. \**p* < .05, \*\**p* < .01, \*\*\**p* < .001

We observed a major impact of increased window sizes on false detections per hour, with a subsequent median reduction from 5.6 FD/h for 1 second, 4.1 FD/h for 2 seconds, 3.2 FD/h for 4 seconds and 2.5 FD/h for 8 seconds. There was a significant difference between 1 second windows and 2 seconds (*β* = −1.31, *p* < .001), 4 seconds (*β* = −2.20, *p* < .001) and 8 seconds (*β* = −2.73, *p* < .001).

With larger windows, the average detection delay increased from a median of 1.6 s in 1 second to 3.9 s in 2 seconds, 7.2 s in 4 seconds and 8.0 s in 8 seconds. There was a statistically significant difference between 1 second and 4 seconds (*β* = 6.41, *p* = .005) and 8 seconds (*β* = 7.27, *p* = .001).

While mean CNN energy consumption was 49.15 µJ/s in 1 second, 41.33 µJ/s in 2 seconds and 36.80 µJ/s in 4 seconds, a window size of 8 seconds exceeded the input size of the CNN accelerator and therefore led to a consumption of 91.34 µJ/s. ADC energy consumption was 3.79 µJ/s for all window sizes.

Compared to non-overlapping windows, using a 50% overlap led to an increase in sensitivity and a decrease in the average detection delay (Figure S1 and Table S14). The CNN energy consumption increased by up to twofold, while the ADC energy consumption stayed the same (Table S4 and S9).

### Impact of digital resolution

When using different digital resolutions (Figure 4), median sensitivity was 83.3% for 16 bits, 80.0% for 14 bits, 82.4% for 12 bits, 75.0% for 10 bits and 69.2% for 8 bits. There was a significant difference between 16 and 8 bits (*β* = −4.22, *p* = .007).

**Figure 4.**
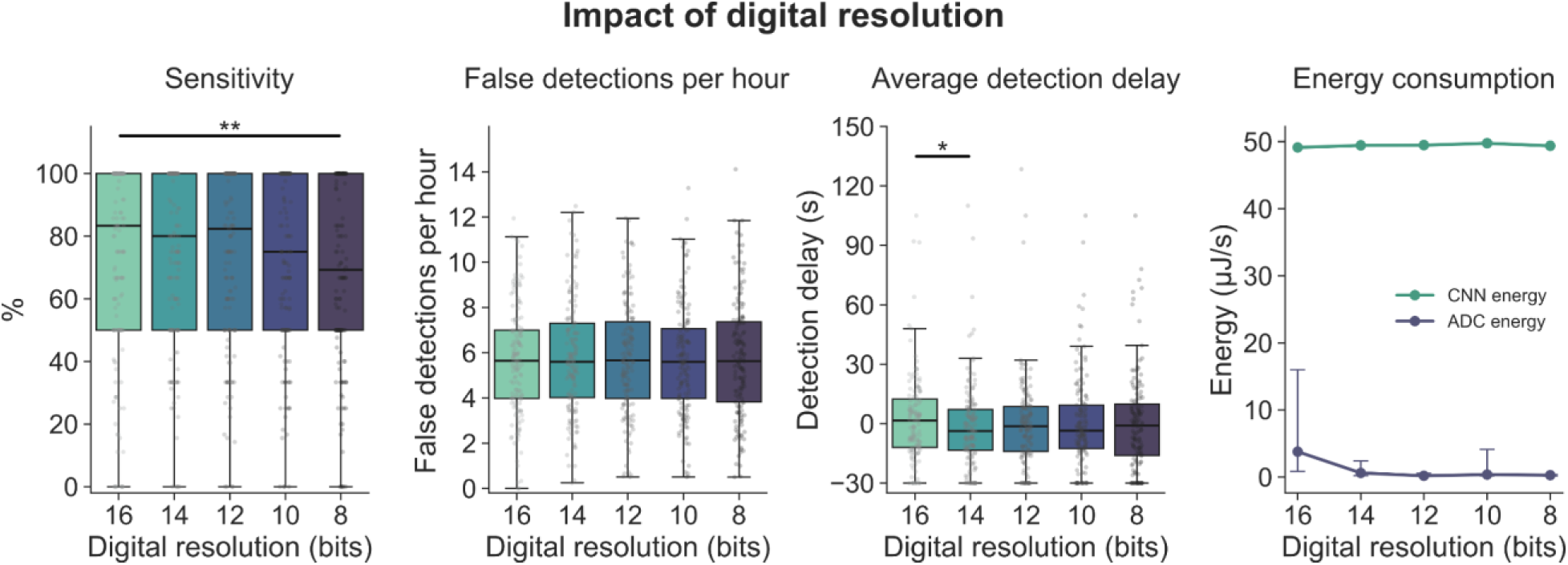
Impact of digital resolution on performance metrics and energy consumption. The gray points represent the individual patients. The error bars for ADC energy consumption represent the IQR. \**p* < .05, \*\**p* < .01

False detections per hour showed median values of 5.6 FD/h for 16, 14, 10 and 8 bits and 5.7 FD/h for 12 bits. There was no significant difference.

When analyzing the average detection delay along decreased digital resolutions, it showed a median of 1.6 s for 16 bits, −3.7 s for 14 bits, −1.3 s for 12 bits, −3.5 s for 10 bits and −1.0 s for 8 bits. There was a significant difference between 16 bits and 14 bits (*β* = −3.90, *p* = .041).

Different digital resolutions only had a negligible impact on mean CNN energy consumption with 49.15 µJ/s for 16 bits, 49.45 µJ/s for 14 bits, 49.49 µJ/s for 12 bits, 49.77 µJ/s for 10 bits and 49.39 µJ/s for 8 bits. The estimated ADC energy showed a consumption of 3.79 µJ/s for 16 bits, 0.61 µJ/s for 14 bits, 0.19 µJ/s for 12 bits, 0.37 µJ/s for 10 bits and 0.28 µJ/s for 8 bits.

### Impact of number of channels

In a final step, we analyzed the impact of the used number of channels on seizure detection performance (Figure 5). Four channels showed a median sensitivity of 100.0%, followed by 85.7% for three channels, 83.3% for two channels and 75.0% for one channel. There was a significant difference between four channels and both two channels (*β* = −8.10, *p* < .001) and one channel (*β* = −9.06, *p* < .001).

**Figure 5.**
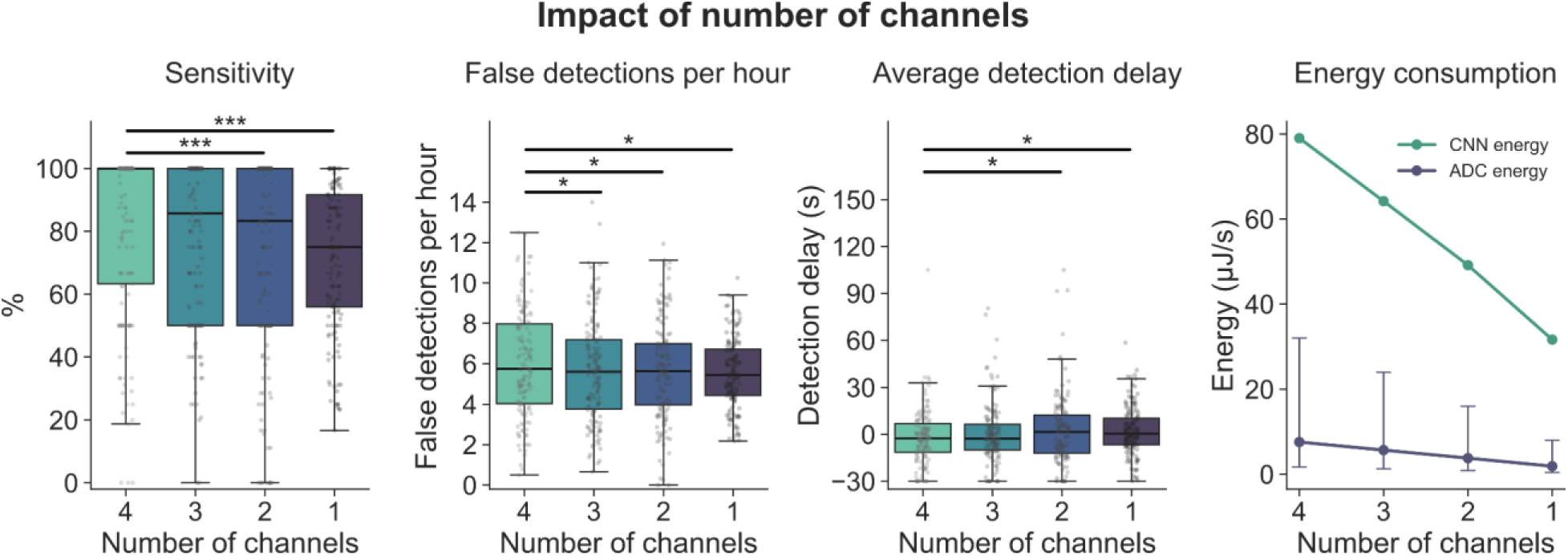
Impact of number of channels on performance metrics and energy consumption. The gray points represent the individual patients. The error bars for ADC energy consumption represent the IQR. \**p* < .05, \*\*\**p* < .001

The false detections per hour showed a median value of 5.8 FD/h for four channels and 5.6 FD/h for three and two channels. Using one channel showed a median of 5.4 FD/h. A significant difference was found when comparing four channels with three channels (*β* = −0.35, *p* = .045), two channels (*β* = −0.41, *p* = .020) and one channel (*β* = −0.49, *p* = .005).

The average detection delay showed a median of −2.5 s when using four channels, −2.8 s for three channels, 1.6 s for two channels and 0.5 s for one channel. There was a significant difference between four channels and both two (*β* = 4.58, *p* = .014) and one channel (*β* = 4.25, *p* = .020).

CNN power consumption was majorly affected by using different numbers of channels, with four channels showing a median of 79.04 µJ/s, followed by 64.22 µJ/s for three channels, 49.15 µJ/s for two channels and 31.63 µJ/s for one channel. This was also the case for ADC energy, where the median consumption was 7.58 µJ/s for four channels, 5.68 µJ/s for three channels, 3.79 µJ/s for two channels and 1.89 µJ/s for one channel.

## Discussion

Fast and reliable seizure detection has the potential to improve the efficacy of all currently available EEG-based neurostimulation devices that treat drug-resistant focal epilepsy, in order to reduce seizure frequency or even completely suppress seizures (10). In the current literature, many approaches were published with a wide range of detection frameworks with varying complexity and preprocessing strategies, primarily evaluated using a small number of patients (17,42). In our work, we address current limitations by 1) using an energy-efficient neural network, 2) systematically investigating the impact of EEG preprocessing parameters and 3) evaluating the performance with a large group of patients.

We chose a patient-specific seizure detection approach because it generally shows better seizure detection performance compared to patient-generic or hybrid approaches. However, a patient-specific approach involves a higher implementation effort (43), since seizure detection models need to be trained with previously collected, patient-specific EEG data, where seizures are either manually marked by experts or automatically marked by seizure detection models (44), which then also need to be validated by experts. Future research should focus on advancing approaches that are not entirely dependent on patient-individual EEG data, since they could reach market readiness faster and thus help patients in a more timely manner.

All seizure detection results are dependent on the chosen classification threshold (45). However, only a limited number of studies report on how the chosen thresholds were determined (46,47). In this study, we report all parameter choices in order to enhance transparency and comparability. We also observed a large interindividual variability in all evaluated seizure detection metrics. One possible reason could be differences in data quality, an issue that has previously been investigated when using wearables for seizure monitoring (48). Data quality can be improved by using a second model for artifact detection (49). However, including more advanced preprocessing techniques usually leads to an increased energy consumption. The evaluation of this tradeoff should be the subject of future research.

Lowering the sampling rate showed a decrease in sensitivity. Interestingly, there was also a decrease in FD/h, which could be due to lower signal complexity. No significant impact on the average detection delay was observed, indicating that this metric is robust to lower sampling rates. When compared to previous studies that used 256 Hz, our results show a substantial improvement in the average detection delay and FD/h (46). However, comparability is limited since the metrics were not calculated as proposed by the SzCORE framework. In a closed-loop neurostimulation approach, lowering the sampling rate can reduce energy consumption by over 60% while maintaining reasonable seizure detection performance.

The metric with the strongest overall impact on seizure detection performance and energy consumption was the chosen window size. When using both non-overlapping and overlapping windows, sensitivity especially decreased when the window size was prolonged to 8 seconds. Larger window sizes also led to a substantial reduction in FD/h, which goes in line with a previous work that compared similar window sizes using sample-based performance metrics (20). In contrast, longer windows significantly increased the average detection delay. A previously proposed model named *SeizureNet* used a window size of 1 second with 50% overlap to obtain a median sensitivity of 96%, 10.1 FD/h and an average detection delay of 3.7 seconds (50). Apart from a slightly higher median sensitivity of 100%, our approach shows substantially lower values regarding FD/h and average detection delay. However, comparability is limited since intracranial EEG data from 22 EPILEPSIAE patients was used. For 8 second windows with and without an overlap, CNN energy consumption was substantially larger, since the maximum CNN accelerator input size of the used microcontroller was exceeded. Using a different microcontroller can resolve this limitation (39). Overall, larger window sizes can especially be useful for applications where FD/h should be minimized, such as wearables that warn the patient on upcoming seizures (31).

Using different digital resolutions only had a small impact on the three seizure detection performance metrics of interest. This can be explained to the applied quantization level of the CNN, where the input data and model weights are always reduced to a digital resolution of 8 bits. A recent study used different CNN quantization levels, where a reduction in performance was only observed when using digital resolutions of less than 8 bits (51). However, the used datasets also included intracranial EEG recordings from non-human subjects, which limits comparability with our work. We observed a significant difference in sensitivity between 16 and 8 bits and in the average detection delay between 16 and 14 bits. This suggests that reducing the digital resolution has to some extent an impact on seizure detection performance. Minor, non-significant fluctuations were observed in the FD/h. Due to the model quantization, the impact on CNN energy consumption was only negligible. In contrast, ADC energy consumption is reduced when using digital resolutions smaller than 16 bits, which has implications on the choice of a suitable ADC for designing a closed-loop neurostimulation device.

Usually, all available EEG channels are used for seizure detection (17,43). It is important to highlight that previous studies used computational approaches to identify optimal channel combinations (52) or systematically reduced the number of channels (53), using standard EEG setups as a basis. However, they used non-CNN approaches and only reported sample-based performance metrics, which limits comparability with our work. Since standard EEG setups with many channels are not practicable for a closed-loop neurostimulation device, studies also explored the use of CNNs with a smaller number of channels and reduced them to up to two channels (33,46). To the best of our knowledge, this work represents the first systematic analysis on the effects of reducing the number of channels from four to only one channel in a CNN-based ultra-low power application. As expected, sensitivity was highest when using four channels and was subsequently reduced when lowering the number of channels. Interestingly, FD/h were minimally reduced when lowering the number of channels and the average detection delay increased when using two or one channel. This could be due to a reduced complexity of the model input. Since we only used channels that cover the seizure onset zone, they are spatially close and thus share mutual information (54). Furthermore, the four channels were always re-referenced to a central channel that was determined in the selection procedure and subsequently reduced afterwards (38). In future studies, the effects of different re-referencing schemes (55) on seizure detection performance should be explored. An important aspect in the choice of the number of channels is power consumption, where we observed a major reduction in CNN and ADC energy when reducing the number of channels. Overall, we confirm and extend the findings of previous studies that a small number of channels is sufficient for performing reliable ultra-low-power seizure detection, provided that they are close to the seizure onset zone.

In the context of closed-loop neurostimulation, systems such as RNS triggered 600 to 2000 stimulations per day in patients which participated in clinical trials (56). Given that our approach showed in individual patients up to 14.1 FD/h, corresponding to 338.4 false detections per day, it would trigger substantially fewer daily stimulations. This also leads to a significant reduction in energy consumption. However, when it is crucial to minimize FD/h, such as in wearables that alert users of potential upcoming seizures (31), further research is needed to reduce FD/h while maintaining low energy consumption.

To our best of knowledge, the microcontroller used in this study has never been used in the literature. A previous study (46) estimated CNN energy consumption on a low-power microcontroller, which does not possess a CNN accelerator. With a window size of 1 second and 4 EEG channels, the model required approximately 243 μJ per detection. For similar conditions, our model required 67% less CNN energy. Another study (30) implemented a CNN on a non-accelerated microcontroller using a 1 second window size and 9 EEG channels. They reported a CNN energy consumption of approximately 215 μJ, which corresponds to 24 µJ/channel. We measured a mean CNN energy of 79 μJ with a non-overlapping window size of 1 second and 4 channels of EEG, which corresponds to 20 μJ/channel. Another limitation of our energy consumption investigation is that only the ADC and CNN energy were considered. Data preprocessing is also expected to have a non-negligible energy cost. However, to the best of our knowledge, the present study is the first work in the context of seizure detection which takes into consideration the ADC energy. This inclusion has shown to be important for the characterization of the impact of digital resolution, because the CNN energy did not vary substantially while the ADC energy was affected.

## Conclusion

This study represents the first systematic analysis on the impact of EEG preprocessing on seizure detection in the context of an ultra-low-power application. We show that seizure detection performance and energy consumption are especially influenced by the chosen window size and number of channels. Our study applies a novel event-based evaluation framework on a large dataset, where we show that seizure detection can be performed with detection delays below one second. We address common issues in the current literature and offer guidance on the selection of appropriate EEG preprocessing parameters for closed-loop neurostimulation approaches. These insights should ultimately support in providing more effective treatment options for patients with drug-resistant focal epilepsy.

## Author contributions

PR: Conceptualization, Data curation, Formal Analysis, Investigation, Methodology, Software, Visualization, Writing – original draft, Writing – review & editing; JL: Data curation, Formal Analysis, Methodology, Resources, Software, Validation, Writing – original draft, Writing – review & editing; CA: Conceptualization, Funding acquisition, Project administration, Supervision, Writing – original draft, Writing – review & editing; SK: Conceptualization, Validation, Writing – review & editing; NZ: Conceptualization, Validation, Writing – review & editing; MD: Data curation, Methodology, Resources, Writing – review & editing; ASB: Funding acquisition, Supervision, Writing – review & editing; PW: Funding acquisition, Supervision, Writing – review & editing; LC: Funding acquisition, Supervision, Writing – review & editing

## Ethical publication statement

We confirm that we have read the Journal’s position on issues involved in ethical publication and affirm that this report is consistent with those guidelines.

## Conflict of interest statement

PR and CA are employees of Precisis GmbH. ASB has received research support and personal honoraria for lectures and advice from Precisis GmbH. The remaining authors have no conflicts of interest.

## Funding statement

This work was supported by the German federal state of Baden-Württemberg, through the Invest BW Project “Brain-MEP” (BW1_1276/03).

## Data availability statement

The data and code to reproduce figures and statistics are available at GitHub (https://github.com/reispat/impact_preproc_seizure_detection).

## Notes

### Funding Statement

This work was supported by the German federal state of Baden-Wuerttemberg, through the Invest BW Project "Brain-MEP" (BW1_1276/03).

### Author Declarations

The Ethics Committees of the three hospitals involved in the development of the EPILEPSIAE database (Ethik-Kommission der Albert-Ludwigs-Universitaet, Freiburg; Comite consultatif sur le traitement de l'information en matiere de recherche dans le domaine de la sante, Hospital Universitario Pitie-Salpetriere; and Ethics Committee of the Centro Hospitalar e Universitario de Coimbra) gave ethical approval for this work.

### Summary of Updates

Table S10 & Figure S1 revised; Supplementary material updated.

