## Supplementary material for "The impact of EEG preprocessing parameters on ultra-low-power seizure detection"

*Details on EEG preprocessing*

EEG data was cropped to 1 hour segments (M = 1.04 h, SD = 0.75 h), including at least one seizure (M = 1.13 seizures, SD = 0.50 seizures), located in the middle of the file. Flat segments were detected and subsequently rejected in the data of 28 patients. The data were filtered with a $5^{th}$ order Butterworth IIR (infinite impulse response) bandpass filter with a high-pass frequency of 0.5 Hz and a low-pass frequency of 48 Hz in order to remove the DC offset and power line noise. The frequency of the low-pass filter was reduced to 30 Hz when using a sampling rate of 64 Hz due to the Nyquist-Shannon sampling theorem.

*Details on the seizure detection model*

The original version of EpiDeNet (1) consists of 5 blocks, each containing a convolutional layer followed by a pooling layer and a ReLU activation function. Although not directly reported in the publication, EpiDeNet also included batch normalization after each convolutional layer, in order to improve speed and stability of the model training. We added additional Dropout layers to prevent overfitting.

The seizure detection models were compiled with an Adam optimizer with a learning rate of $1e^{-4}$ and a binary cross-entropy loss function. Since the data in seizure detection problems is usually heavily imbalanced (2) and only 5.8% of the data in the used dataset are labeled as a seizure, class weighting was applied to the data prior to model training using scikit-learn (3). The models were then trained with a fixed number of 300 epochs, a batch size of 512 and 20% of the training data were used for validation. Model training was accelerated with an NVIDIA A2 GPU using CUDA. For each model, random seeds were kept constant to ensure reproducibility.

Ultra-low-power seizure detection is usually performed on a microcontroller, thus we have converted and optimized the models using TensorFlow Lite (TFLite), integrated in TensorFlow. Specifically, the model weights were quantized to 8-bit integers using full integer quantization so that the minimum and maximum values correspond to -128 and 127 respectively. The quantization was conducted using the training data as a representative dataset. A quantize layer was added before the first layer of the model to convert the 32-bit floating point EEG data to 8-bit integers. Similarly, a dequantize layer was added after the last layer of the model to convert the 8-bit integer model output to 32-bit floating point.

*Details on sampling rate*

Downsampling was used to approximate an input signal with a lower sampling frequency. To avoid aliasing, the high-frequency components of the signal were removed using a low-pass filter. Afterwards, the signal was decimated to obtain the required number of samples per second. When the target frequency was 1/X of the original frequency, the decimation consisted in retaining every $X^{th}$ sample (4). This procedure was done using the function *resample* from MNE-Python.

*Details on window size*

The windowing was done using the function *make_fixed_length_epochs* from MNE-Python. Increasing the window length usually leads to increases in CNN energy consumption and the average detection delay. To prevent the latter effect, overlapping windows were also investigated. For example, with a window of 4 seconds and a 50% overlap, a detection is performed every 2 seconds, including data from the past 4 seconds.

*Details on digital resolution*

To simulate the impacts of a lower resolution ADC, the EEG floating point values were first converted back to integer values with the known scaling factor. These integer values were then right-shifted to decrease the resolution and converted back to floating-point values with a new scaling factor. The complete procedure is shown in the table S1.

| Step | Procedure |
|---|---|
| | ReduceSamplingResolution(original_eeg, original_scaling_factor, original_resolution, new_resolution) |
| 1 | original_eeg_int ← original_eeg / scaling_factor |
| 2 | Right_shift ← original_resolution - new_resolution |
| 3 | New_eeg_int ← original_eeg_int >> right_shift |
| 4 | New_scaling_factor ← original_scaling_factor * 2**right_shift |

| | |
|---|---|
| 5 | New_eeg ← new_eeg_int * new_scaling_factor |
| 6 | Return new_eeg |
| | End Procedure |

Table S1. Procedure to reduce the digital resolution.

*Details on number of channels*

The EEG recordings consist of a mixture of 10-20 and 10-10 channel montages with 19 to 37 EEG channels. Since the focus of the current approach is on a low-channel application using channels covering the seizure onset zone (SOZ), the number of channels was reduced to one central electrode and four neighboring channels (minimum distance: 40 mm) according to seizure annotations in the corresponding channels across all data of a patient. Afterwards, the neighboring channels were re-referenced to the central channel, leading to a final number of four channels. Further details on the used approach can be found in Manzouri et al. (5) under “Method 2”.

*Details on seizure detection performance*

In the used Seizure Community Open-Source Research Evaluation framework (SzCORE), consecutive time windows of detected seizures are treated as events, with several defined tolerances and overlaps (6). When an event was detected, we allowed any overlap with the true event to be considered as a proper detection (termed *Minimum overlap* in the SzCORE framework). An event was counted as a true detection when it was detected up to 30 seconds before its real occurrence (*Pre-ictal tolerance*). We also considered events that were detected after up to 60 seconds of the real occurrence as correct detections (*Post-ictal tolerance*). When two detected events were less than 90 seconds apart, they were merged into a single event (*Minimum duration*). When an event exceeded five minutes, the event was split into separate events with a maximum length of five minutes (*Maximum event duration*). This was always done after the previously described merging of events. As advised by SzCORE, splitting of such events should be performed because seizures lasting longer than five minutes are considered status epilepticus (6). However, this is only the case in convulsive tonic-clonic status epilepticus (7).

For calculation of the seizure detection performance metrics, we used the timescoring library as provided by the SzCORE framework (https://github.com/esl-epfl/timescoring) and custom tools (https://github.com/HKA-IES/brainmep-nas). All performance metrics are dependent on the chosen classification threshold when an event is counted as a detection. This threshold is usually set to 50% in a balanced two-class problem (8). In the context of seizure detection, where the two classes are normally heavily imbalanced, other strategies are used to set a threshold, such as choosing the threshold that maximizes the F1 score of the currently evaluated testing data (9). Here, we have set thresholds that maximized the F1 score of the training data instead of the testing data, so that the performance on the test set is not artificially enhanced.

We primarily focused on three event-based metrics that are most relevant for an ultra-low-power closed-loop application. Sensitivity, also known as recall, is the ability of the system to identify true seizure events and is usually expressed in percent. Another important metric is the number of false detections per hour (FD/h), in order to avoid additional energy consumption through unnecessary stimulation. In the context of neurostimulation, the time until a seizure has been detected is of major importance. This detection delay was expressed as an average delay of detected events in seconds, since a segment could have included more than one true seizure. The average detection delay could also be negative, since we allowed a pre-ictal tolerance of 30 seconds.

*Details on CNN energy consumption*

The Silicon Labs EFR32MG24 microcontroller is especially suited for a closed-loop application because of the included radio transceiver module supporting Bluetooth, very low sleep mode current, and a matrix vector processor (MVP) for CNN acceleration. The MVP is optimized to perform matrix floating point multiplications and additions, enabling up to 82x faster matrix multiplication for up to 60x lower energy compared to software-only operations (10). Silicon Labs officially supports the deployment of CNNs with TensorFlow Lite (TFLite) for microcontrollers. The CNN model was first developed using TensorFlow and Keras, trained on a computer, and then quantized to 8-bit integers. It was then converted to a Flatbuffer format which can be interpreted by the TFLite runtime on the microcontroller. CNN energy was measured using an automated testbench. Models were automatically converted to C code, compiled, and flashed to the microcontroller with a Raspberry Pi 4B (Raspberry Pi, Cambridge, England). The energy measurement was conducted with a Joulescope JS220 (Jetperch, Olney, USA) and a Voltcraft PPS-16005 voltage source. A Yepkit YKUSH XS USB switch (Yepkit, Alverca, Portugal) was used to temporarily interrupt the USB connection between the Raspberry

Pi and the microcontroller, to ensure that the energy measurements are realized without external influences. The energy measurements were conducted with a supply voltage of 1.8 V and a frequency of 78 MHz. Measurements were repeated five times. The reported flash memory, RAM, operations, and multiply-accumulates (MACs) are estimated with the Silicon Labs MLTK Model Profiler (https://siliconlabs.github.io/mltk/docs/guides/model_profiler.html).

| Sampling rate | CNN energy per inference (Mean) | CNN energy per second (Mean) | CNN latency (Mean) | Flash memory | RAM | Trainable parameters | Operations | MACs |
|---|---|---|---|---|---|---|---|---|
| 256 Hz | 49.15 µJ | 49.15 µJ/s | 3.92 ms | 17.4 kB | 6.9 kB | 9453 | 269.7k | 127.0k |
| 128 Hz | 29.41 µJ | 29.41 µJ/s | 2.39 ms | 17.4 kB | 5.5 kB | 9453 | 134.9k | 63.5k |
| 64 Hz | 17.26 µJ | 17.26 µJ/s | 1.41 ms | 11.1 kB | 5.4 kB | 3741 | 54.0k | 25.1k |

Table S2. CNN energy consumption and additional parameters for the analyzed sampling rates.

| Window size | CNN energy per inference (Mean) | CNN energy per second (Mean) | CNN latency (Mean) | Flash memory | RAM | Trainable parameters | Operations | MACs |
|---|---|---|---|---|---|---|---|---|
| 1 s | 49.15 µJ | 49.15 µJ/s | 3.92 ms | 17.4 kB | 6.9 kB | 9453 | 269.7k | 127.0k |
| 2 s | 82.66 µJ | 41.33 µJ/s | 6.86 ms | 17.4 kB | 10.5 kB | 9453 | 539.4k | 254.0k |
| 4 s | 147.20 µJ | 36.80 µJ/s | 12.63 ms | 17.4 kB | 17.7 kB | 9453 | 1.1M | 507.9k |
| 8 s | 731.0 µJ | 91.34 µJ/s | 60.11 ms | 17.4 kB | 23.9 kB | 9453 | 2.2M | 1.0M |

Table S3. CNN energy consumption and additional parameters for the analyzed window sizes (0% overlap).

| Window size | CNN energy per inference (Mean) | CNN energy per second (Mean) | CNN latency (Mean) | Flash memory | RAM | Trainable parameters | Operations | MACs |
|---|---|---|---|---|---|---|---|---|
| 1 s | 49.19 µJ | 98.39 µJ/s | 3.92 ms | 17.4 kB | 6.9 kB | 9453 | 269.7k | 127.0k |
| 2 s | 82.40 µJ | 82.39 µJ/s | 6.86 ms | 17.4 kB | 10.5 kB | 9453 | 539.4k | 254.0k |
| 4 s | 144.90 µJ | 72.45 µJ/s | 12.63 ms | 17.4 kB | 17.7 kB | 9453 | 1.1M | 507.9k |

| | | | | | | | | |
|---|---|---|---|---|---|---|---|---|
| 8 s | 730.90 µJ | 182.73 µJ/s | 60.11 ms | 17.4 kB | 23.9 kB | 9453 | 2.2M | 1.0M |

Table S4. CNN energy consumption and additional parameters for the analyzed window sizes (50% overlap).

| Digital resolution | CNN energy per inference (Mean) | CNN energy per second (Mean) | CNN latency (Mean) | Flash memory | RAM | Trainable parameters | Operations | MACs |
|---|---|---|---|---|---|---|---|---|
| 16 bits | 49.15 µJ | 49.15 µJ/s | 3.92 ms | 17.4 kB | 6.9 kB | 9453 | 269.7k | 127.0k |
| 14 bits | 49.45 µJ | 49.45 µJ/s | 3.92 ms | 17.4 kB | 6.9 kB | 9453 | 269.7k | 127.0k |
| 12 bits | 49.49 µJ | 49.49 µJ/s | 3.92 ms | 17.4 kB | 6.9 kB | 9453 | 269.7k | 127.0k |
| 10 bits | 49.77 µJ | 49.77 µJ/s | 3.92 ms | 17.4 kB | 6.9 kB | 9453 | 269.7k | 127.0k |
| 8 bits | 49.39 µJ | 49.39 µJ/s | 3.92 ms | 17.4 kB | 6.9 kB | 9453 | 269.7k | 127.0k |

Table S5. CNN energy consumption and additional parameters for the analyzed digital resolutions.

| Number of channels | CNN energy per inference (Mean) | CNN energy per second (Mean) | CNN latency (Mean) | Flash memory | RAM | Trainable parameters | Operations | MACs |
|---|---|---|---|---|---|---|---|---|
| 1 channel | 31.63 µJ | 31.63 µJ/s | 2.59 ms | 17.4 kB | 5.5 kB | 9453 | 139.1k | 65.6k |
| 2 channels | 49.15 µJ | 49.15 µJ/s | 3.92 ms | 17.4 kB | 6.9 kB | 9453 | 269.7k | 127.0k |
| 3 channels | 64.22 µJ | 64.22 µJ/s | 5.23 ms | 17.4 kB | 8.7 kB | 9453 | 400.4k | 188.4k |
| 4 channels | 79.04 µJ | 79.04 µJ/s | 6.57 ms | 17.4 kB | 10.5 kB | 9453 | 531.1k | 249.9k |

Table S6. CNN energy consumption and additional parameters for the analyzed numbers of channels.

*Details on ADC energy consumption*

Instead of extrapolating ADC energy measurements from the data sheets of a few selected commercially available components, we used Boris Murmann's ADC Performance Survey (11). We considered only delta-sigma ADCs. The ADCs resolution is not directly reported, however, the effective number of bits (ENOB) is reported in the survey and corresponds to the real resolution, taking into account noise and distortion. We assumed that ENOB is 2 bits lower than the desired digital resolution. We have grouped the ADCs in ranges of 2 bits and calculated the median as well as the 25th and 75th percentiles, which are reported as the interquartile range (IQR). These three values are expected to give a good indication of the performance of commercially available ADCs. The estimated ADC sample energy for each digital resolution is shown in the table below.

| Digital Resolution | Approx. ENOB | 25$^{th}$ percentile | Median | 75$^{th}$ percentile |
|---|---|---|---|---|
| 8 bits | 6 ± 1 bits | 303 pJ | 543 pJ | 782 pJ |
| 10 bits | 8 ± 1 bits | 338 pJ | 725 pJ | 8065 pJ |
| 12 bits | 10 ± 1 bits | 214 pJ | 378 pJ | 1069 pJ |
| 14 bits | 12 ± 1 bits | 388 pJ | 1182 pJ | 4678 pJ |
| 16 bits | 14 ± 1 bits | 1667 pJ | 7400 pJ | 31250 pJ |

Table S7. Estimated ADC sample energy for each digital resolution.

| | ADC energy per inference | | | ADC energy per second | | |
|---|---|---|---|---|---|---|
| Sampling rate | 25$^{th}$ percentile | Median | 75$^{th}$ percentile | 25$^{th}$ percentile | Median | 75$^{th}$ percentile |
| 256 Hz | 0.85 μJ | 3.79 μJ | 16 μJ | 0.85 μJ/s | 3.79 μJ/s | 16 μJ/s |
| 128 Hz | 0.43 μJ | 1.89 μJ | 8 μJ | 0.43 μJ/s | 1.89 μJ/s | 8 μJ/s |
| 64 Hz | 0.21 μJ | 0.95 μJ | 4 μJ | 0.21 μJ/s | 0.95 μJ/s | 4 μJ/s |

Table S8. ADC energy consumption for the analyzed sampling rates.

| | ADC energy per inference | | | ADC energy per second | | |
|---|---|---|---|---|---|---|
| Window size | 25$^{th}$ percentile | Median | 75$^{th}$ percentile | 25$^{th}$ percentile | Median | 75$^{th}$ percentile |
| 1 s | 0.85 μJ | 3.79 μJ | 16 μJ | 0.85 μJ/s | 3.79 μJ/s | 16 μJ/s |
| 2 s | 1.71 μJ | 7.58 μJ | 32 μJ | 0.85 μJ/s | 3.79 μJ/s | 16 μJ/s |
| 4 s | 3.41 μJ | 15.16 μJ | 64 μJ | 0.85 μJ/s | 3.79 μJ/s | 16 μJ/s |
| 8 s | 6.83 μJ | 30.31 μJ | 128 μJ | 0.85 μJ/s | 3.79 μJ/s | 16 μJ/s |

Table S9. ADC energy consumption for the analyzed window sizes (0% overlap).

| | ADC energy per inference | | | ADC energy per second | | |
|---|---|---|---|---|---|---|
| Window size | 25th percentile | Median | 75th percentile | 25th percentile | Median | 75th percentile |
| 1 s | 0.85 µJ | 3.79 µJ | 16 µJ | 0.85 µJ/s | 3.79 µJ/s | 16 µJ/s |
| 2 s | 1.71 µJ | 7.58 µJ | 32 µJ | 0.85 µJ/s | 3.79 µJ/s | 16 µJ/s |
| 4 s | 3.41 µJ | 15.16 µJ | 64 µJ | 0.85 µJ/s | 3.79 µJ/s | 16 µJ/s |
| 8 s | 6.83 µJ | 30.31 µJ | 128 µJ | 0.85 µJ/s | 3.79 µJ/s | 16 µJ/s |

Table S10. ADC energy consumption for the analyzed window sizes (50% overlap).

| | ADC energy per inference | | | ADC energy per second | | |
|---|---|---|---|---|---|---|
| Digital resolution | 25th percentile | Median | 75th percentile | 25th percentile | Median | 75th percentile |
| 16 bits | 0.85 µJ | 3.79 µJ | 16 µJ | 0.85 µJ/s | 3.79 µJ/s | 16 µJ/s |
| 14 bits | 0.20 µJ | 0.61 µJ | 2.40 µJ | 0.20 µJ/s | 0.61 µJ/s | 2.40 µJ/s |
| 12 bits | 0.11 µJ | 0.19 µJ | 0.55 µJ | 0.11 µJ/s | 0.19 µJ/s | 0.55 µJ/s |
| 10 bits | 0.17 µJ | 0.37 µJ | 4.13 µJ | 0.17 µJ/s | 0.37 µJ/s | 4.13 µJ/s |
| 8 bits | 0.16 µJ | 0.28 µJ | 0.40 µJ | 0.16 µJ/s | 0.28 µJ/s | 0.40 µJ/s |

Table S11. ADC energy consumption for the analyzed digital resolutions.

| | ADC energy per inference | | | ADC energy per second | | |
|---|---|---|---|---|---|---|
| Number of channels | 25th percentile | Median | 75th percentile | 25th percentile | Median | 75th percentile |
| 4 channels | 1.71 µJ | 7.58 µJ | 32 µJ | 1.71 µJ/s | 7.58 µJ/s | 32 µJ/s |
| 3 channels | 1.28 µJ | 5.68 µJ | 24 µJ | 1.28 µJ/s | 5.68 µJ/s | 24 µJ/s |
| 2 channels | 0.85 µJ | 3.79 µJ | 16 µJ | 0.85 µJ/s | 3.79 µJ/s | 16 µJ/s |
| 1 channels | 0.43 µJ | 1.89 µJ | 8 µJ | 0.43 µJ/s | 1.89 µJ/s | 8 µJ/s |

Table S12. ADC energy consumption for the analyzed numbers of channels.

*Details on statistical analysis*

Across all investigated parameters, the general linear mixed-effects model (LMM) formula was *performance ~ 1 + parameter + (1 | patient)*. The random intercept for patients was added to account for their inherent variability and repeated measures. To perform comparisons between all conditions, post-hoc contrasts were calculated using the emmeans package. To account for multiple comparisons, *p* values were corrected using the Benjamini-Hochberg procedure. For each calculated contrast, we report *p* values and Cohen's *d* as effect size. Descriptives and detailed statistical metrics for the LMMs and post-hoc tests are reported in the following tables.

| *Sensitivity* | | | | | |
|---|---|---|---|---|---|
| Descriptives | | | | | |
| Sampling rate | Mean | SD | Median | | |
| 256 Hz | 72.2% | 31.1% | 83.3% | | |
| 128 Hz | 66.5% | 30.2% | 72.7% | | |
| 64 Hz | 67.0% | 28.8% | 66.7% | | |
| Linear mixed-effects model | | | | | |
| Sampling rate | *β* | *SE* | df | *t* ratio | *p* |
| 128 Hz | -5.70 | 2.12 | 288 | -2.69 | .008* |
| 64 Hz | -5.16 | 2.12 | 288 | -2.44 | .015* |
| Conditional $R^2$: 0.64; Marginal $R^2$: 0.01 | | | | | |
| Post-hoc contrasts | | | | | |
| Contrast | *p* | *d* | | | |
| 256 vs. 128 Hz | .023* | 0.32 | | | |
| 256 vs. 64 Hz | .023* | 0.29 | | | |
| 128 vs. 64 Hz | .799 | -0.03 | | | |

| *False detections per hour* | | | |
|---|---|---|---|
| Descriptives | | | |
| Sampling rate | Mean | SD | Median |
| 256 Hz | 5.6 FD/h | 2.5 FD/h | 5.6 FD/h |
| 128 Hz | 5.3 FD/h | 2.4 FD/h | 5.4 FD/h |
| 64 Hz | 5.0 FD/h | 2.6 FD/h | 4.4 FD/h |
| Linear mixed-effects model | | | |

| Sampling rate | *β* | *SE* | df | *t* ratio | *p* |
|---|---|---|---|---|---|
| 128 Hz | -0.30 | 0.20 | 288 | -1.47 | .143 |
| 64 Hz | -0.64 | 0.20 | 288 | -3.14 | .002* |
| Conditional $R^2$: 0.52; Marginal $R^2$: 0.01 | | | | | |
| Post-hoc contrasts | | | | | |
| Contrast | *p* | *d* | | | |
| 256 vs. 128 Hz | .143 | 0.17 | | | |
| 256 vs. 64 Hz | .006* | 0.37 | | | |
| 128 vs. 64 Hz | .143 | 0.20 | | | |
| | | *Average detection delay* | | | |
| Descriptives | | | | | |
| Sampling rate | Mean | SD | Median | | |
| 256 Hz | 2.5 s | 22.6 s | 1.6 s | | |
| 128 Hz | 1.1 s | 24.2 s | -0.3 s | | |
| 64 Hz | 4.8 s | 26.2 s | 3.0 s | | |
| Linear mixed-effects model | | | | | |
| Sampling rate | *β* | *SE* | df | *t* ratio | *p* |
| 128 Hz | -0.96 | 2.20 | 271 | -0.44 | .663 |
| 64 Hz | 2.78 | 2.20 | 272 | 1.26 | .208 |
| Conditional $R^2$: 0.45; Marginal $R^2$: 0.004 | | | | | |
| Post-hoc contrasts | | | | | |
| Contrast | *p* | *d* | | | |
| 256 vs. 128 Hz | .663 | 0.05 | | | |
| 256 vs. 64 Hz | .312 | -0.15 | | | |
| 128 vs. 64 Hz | .270 | -0.21 | | | |

Table S13. Metrics on the impact of sampling rate. *$p < .05$

| *Sensitivity* | | | | | |
|---|---|---|---|---|---|
| Descriptives | | | | | |
| Window size | Mean | SD | Median | | |
| 1 s | 72.2% | 31.1% | 83.3% | | |
| 2 s | 70.9% | 28.7% | 75.0% | | |
| 4 s | 71.4% | 30.3% | 80.0% | | |
| 8 s | 68.1% | 30.4% | 66.7% | | |
| Linear mixed-effects model | | | | | |
| Window size | $\beta$ | *SE* | df | *t* ratio | *p* |
| 2 s | -1.27 | 1.90 | 432 | -0.67 | .503 |
| 4 s | -0.77 | 1.90 | 432 | -0.41 | .684 |
| 8 s | -4.06 | 1.90 | 432 | -2.14 | .033* |
| Conditional $R^2$: 0.71; Marginal $R^2$: 0.003 | | | | | |
| Post-hoc contrasts | | | | | |
| Contrast | *p* | *d* | | | |
| 1 vs. 2 s | .754 | 0.08 | | | |
| 1 vs. 4 s | .793 | 0.05 | | | |
| 1 vs. 8 s | .198 | 0.25 | | | |
| 2 vs. 4 s | .793 | -0.03 | | | |
| 2 vs. 8 s | .285 | 0.17 | | | |
| 4 vs. 8 s | .252 | 0.20 | | | |
| *False detections per hour* | | | | | |
| Descriptives | | | | | |
| Window size | Mean | SD | Median | | |
| 1 s | 5.6 FD/h | 2.5 FD/h | 5.6 FD/h | | |
| 2 s | 4.3 FD/h | 2.1 FD/h | 4.1 FD/h | | |
| 4 s | 3.4 FD/h | 2.0 FD/h | 3.2 FD/h | | |
| 8 s | 2.9 FD/h | 2.0 FD/h | 2.5 FD/h | | |

| | | | | | |
|---|---|---|---|---|---|
| Linear mixed-effects model | | | | | |
| Window size | *β* | *SE* | df | *t* ratio | *p* |
| 2 s | -1.31 | 0.18 | 432 | -7.13 | $4.14e^{-12}$* |
| 4 s | -2.20 | 0.18 | 432 | -12.02 | < $2.00e^{-16}$* |
| 8 s | -2.73 | 0.18 | 432 | -14.90 | < $2.00e^{-16}$* |
| Conditional $R^2$: 0.57; Marginal $R^2$: 0.19 | | | | | |
| Post-hoc contrasts | | | | | |
| Contrast | *p* | *d* | | | |
| 1 vs. 2 s | < .0001* | 0.84 | | | |
| 1 vs. 4 s | < .0001* | 1.41 | | | |
| 1 vs. 8 s | < .0001* | 1.75 | | | |
| 2 vs. 4 s | < .0001* | 0.57 | | | |
| 2 vs. 8 s | < .0001* | 0.91 | | | |
| 4 vs. 8 s | .004* | 0.34 | | | |
| | | | *Average detection delay* | | |
| Window size | Mean | SD | Median | | |
| 1 s | 2.5 s | 22.6 s | 1.6 s | | |
| 2 s | 2.5 s | 18.2 s | 3.9 s | | |
| 4 s | 9.0 s | 21.7 s | 7.2 s | | |
| 8 s | 9.8 s | 24.7 s | 8.0 s | | |
| Linear mixed-effects model | | | | | |
| Window size | *β* | *SE* | df | *t* ratio | *p* |
| 2 s | -0.07 | 2.26 | 412 | -0.03 | .976 |
| 4 s | 6.41 | 2.27 | 413 | 2.82 | .005* |
| 8 s | 7.27 | 2.26 | 412 | 3.21 | .001* |
| Conditional $R^2$: 0.28; Marginal $R^2$: 0.02 | | | | | |
| Post-hoc contrasts | | | | | |
| Contrast | *p* | *d* | | | |

| | | |
|---|---|---|
| 1 vs. 2 s | .976 | 0.004 |
| 1 vs. 4 s | .008* | -0.34 |
| 1 vs. 8 s | .004* | -0.39 |
| 2 vs. 4 s | .008* | -0.34 |
| 2 vs. 8 s | .004* | -0.39 |
| 4 vs. 8 s | .844 | -0.05 |

Table S14. Metrics on the impact of window size (0% overlap). **p* < .05

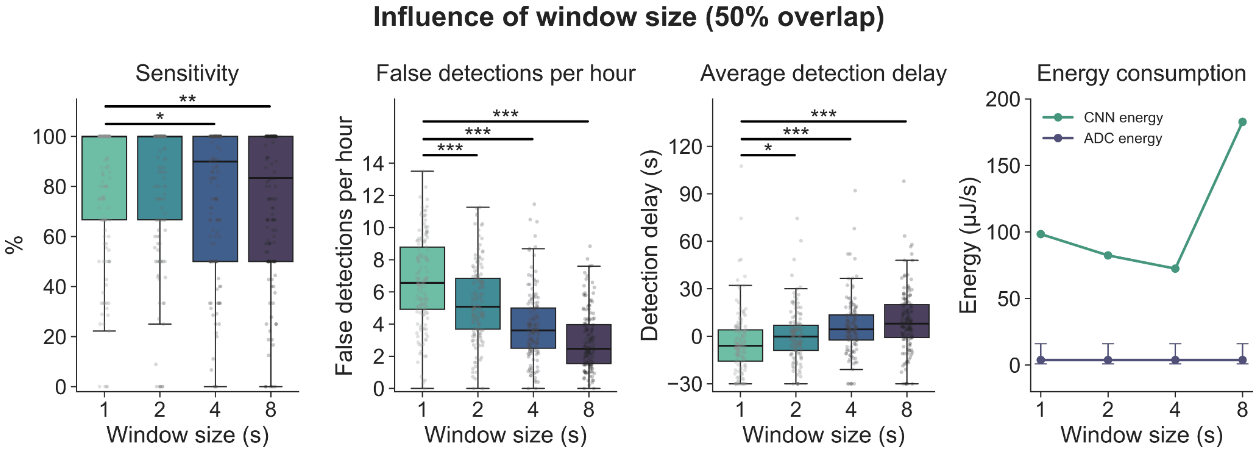


Figure S1. Impact of window size with 50% overlap on performance metrics and energy consumption. The gray points represent the individual patients. The error bars for ADC energy consumption represent the IQR. **p* < .05, ***p* < .01, ****p* < .001

*Sensitivity*

| Descriptives | | | | | |
|---|---|---|---|---|---|
| Window size | Mean | SD | Median | | |
| 1 s | 79.9% | 26.6% | 100.0% | | |
| 2 s | 80.7% | 27.2% | 100.0% | | |
| 4 s | 76.5% | 28.0% | 90.0% | | |
| 8 s | 74.4% | 29.0% | 83.3% | | |
| Linear mixed-effects model | | | | | |
| Window size | *β* | *SE* | df | *t* ratio | *p* |

| | | | | | |
|---|---|---|---|---|---|
| 2 s | 0.77 | 1.68 | 432 | 0.46 | .648 |
| 4 s | -3.37 | 1.68 | 432 | -2.00 | .046* |
| 8 s | -5.51 | 1.68 | 432 | -3.28 | .001* |
| Conditional $R^2$: 0.74; Marginal $R^2$: 0.01 | | | | | |
| Post-hoc contrasts | | | | | |
| Contrast | *p* | *d* | | | |
| 1 vs. 2 s | .648 | -0.05 | | | |
| 1 vs. 4 s | .069 | 0.24 | | | |
| 1 vs. 8 s | .003* | 0.39 | | | |
| 2 vs. 4 s | .029* | 0.29 | | | |
| 2 vs. 8 s | .001* | 0.44 | | | |
| 4 vs. 8 s | .244 | 0.15 | | | |
| | | | *False detections per hour* | | |
| Descriptives | | | | | |
| Window size | Mean | SD | Median | | |
| 1 s | 6.6 FD/h | 2.8 FD/h | 6.6 FD/h | | |
| 2 s | 5.4 FD/h | 2.4 FD/h | 5.1 FD/h | | |
| 4 s | 3.9 FD/h | 2.2 FD/h | 3.6 FD/h | | |
| 8 s | 3.0 FD/h | 1.8 FD/h | 2.5 FD/h | | |
| Linear mixed-effects model | | | | | |
| Window size | *β* | *SE* | df | *t* ratio | *p* |
| 2 s | -1.27 | 0.17 | 432 | -7.29 | $1.46e^{-12}$* |
| 4 s | -2.76 | 0.17 | 432 | -15.86 | < $2.00e^{-16}$* |
| 8 s | -3.66 | 0.17 | 432 | -21.01 | < $2.00e^{-16}$* |
| Conditional $R^2$: 0.70; Marginal $R^2$: 0.27 | | | | | |
| Post-hoc contrasts | | | | | |
| Contrast | *p* | *d* | | | |
| 1 vs. 2 s | < .0001* | 0.86 | | | |

| | | | | | |
|---|---|---|---|---|---|
| 1 vs. 4 s | < .0001* | 1.86 | | | |
| 1 vs. 8 s | < .0001* | 2.47 | | | |
| 2 vs. 4 s | < .0001* | 1.01 | | | |
| 2 vs. 8 s | < .0001* | 1.61 | | | |
| 4 vs. 8 s | < .0001* | 0.61 | | | |
| *Average detection delay* | | | | | |
| Descriptives | | | | | |
| Window size | Mean | SD | Median | | |
| 1 s | -4.0 s | 20.1 s | -6.0 s | | |
| 2 s | 0.2 s | 16.6 s | -0.2 s | | |
| 4 s | 5.8 s | 17.8 s | 4.3 s | | |
| 8 s | 9.4 s | 19.7 s | 8.0 s | | |
| Linear mixed-effects model | | | | | |
| Window size | $\beta$ | *SE* | df | *t* ratio | *p* |
| 2 s | 4.21 | 1.73 | 411 | 2.44 | .015* |
| 4 s | 9.58 | 1.72 | 411 | 5.58 | $4.34e^{-08}$* |
| 8 s | 13.04 | 1.72 | 412 | 7.57 | $2.55e^{-13}$* |
| Conditional $R^2$: 0.45; Marginal $R^2$: 0.07 | | | | | |
| Post-hoc contrasts | | | | | |
| Contrast | *p* | *d* | | | |
| 1 vs. 2 s | .018* | -0.29 | | | |
| 1 vs. 4 s | < .0001* | -0.66 | | | |
| 1 vs. 8 s | < .0001* | -0.91 | | | |
| 2 vs. 4 s | .003* | -0.37 | | | |
| 2 vs. 8 s | < .0001* | -0.61 | | | |
| 4 vs. 8 s | .045* | -0.24 | | | |

Table S15. Metrics on the impact of window size (50% overlap). *$p < .05$

| | | *Sensitivity* | | | |
|---|---|---|---|---|---|
| Descriptives | | | | | |
| Digital resolution | Mean | SD | Median | | |
| 16 bits | 72.2% | 31.1% | 83.3% | | |
| 14 bits | 73.4% | 28.2% | 80.0% | | |
| 12 bits | 73.9% | 28.1% | 82.4% | | |
| 10 bits | 71.5% | 28.5% | 75.0% | | |
| 8 bits | 67.9% | 29.8% | 69.2% | | |
| Linear mixed-effects model | | | | | |
| Digital resolution | $\beta$ | *SE* | df | *t* ratio | *p* |
| 14 bits | 1.27 | 1.57 | 576 | 0.81 | .416 |
| 12 bits | 1.71 | 1.57 | 576 | 1.09 | .275 |
| 10 bits | -0.68 | 1.57 | 576 | -0.43 | .666 |
| 8 bits | -4.22 | 1.57 | 576 | -2.70 | .007* |
| Conditional $R^2$: 0.79; Marginal $R^2$: 0.01 | | | | | |
| Post-hoc contrasts | | | | | |
| Contrast | *p* | *d* | | | |
| 16 vs. 14 bits | .52 | -0.10 | | | |
| 16 vs. 12 bits | .393 | -0.13 | | | |
| 16 vs. 10 bits | .74 | 0.05 | | | |
| 16 vs. 8 bits | .024* | 0.32 | | | |
| 14 vs. 12 bits | .78 | -0.03 | | | |
| 14 vs. 10 bits | .356 | 0.15 | | | |
| 14 vs. 8 bits | .002* | 0.41 | | | |
| 12 vs. 10 bits | .256 | 0.18 | | | |
| 12 vs. 8 bits | .002* | 0.45 | | | |
| 10 vs. 8 bits | .06 | 0.27 | | | |

| *False detections per hour* | | | | | |
|---|---|---|---|---|---|
| Descriptives | | | | | |
| Digital resolution | Mean | SD | Median | | |
| 16 bits | 5.6 FD/h | 2.5 FD/h | 5.6 FD/h | | |
| 14 bits | 5.8 FD/h | 2.5 FD/h | 5.6 FD/h | | |
| 12 bits | 5.8 FD/h | 2.5 FD/h | 5.7 FD/h | | |
| 10 bits | 5.7 FD/h | 2.5 FD/h | 5.6 FD/h | | |
| 8 bits | 5.7 FD/h | 2.6 FD/h | 5.6 FD/h | | |
| Linear mixed-effects model | | | | | |
| Digital resolution | *β* | *SE* | df | *t* ratio | *p* |
| 14 bits | 0.15 | 0.17 | 576 | 0.89 | .374 |
| 12 bits | 0.14 | 0.17 | 576 | 0.82 | .414 |
| 10 bits | 0.04 | 0.17 | 576 | 0.24 | .809 |
| 8 bits | 0.13 | 0.17 | 576 | 0.76 | .447 |
| Conditional $R^2$: 0.67; Marginal $R^2$: 0.001 | | | | | |
| Post-hoc contrasts | | | | | |
| Contrast | *p* | *d* | | | |
| 16 vs. 14 bits | .955 | -0.10 | | | |
| 16 vs. 12 bits | .955 | -0.10 | | | |
| 16 vs. 10 bits | .955 | -0.03 | | | |
| 16 vs. 8 bits | .955 | -0.09 | | | |
| 14 vs. 12 bits | .955 | 0.01 | | | |
| 14 vs. 10 bits | .955 | 0.08 | | | |
| 14 vs. 8 bits | .955 | 0.02 | | | |
| 12 vs. 10 bits | .955 | 0.07 | | | |
| 12 vs. 8 bits | .955 | 0.01 | | | |
| 10 vs. 8 bits | .955 | -0.06 | | | |
| *Average detection delay* | | | | | |

| Descriptives | | | | | |
|---|---|---|---|---|---|
| Digital resolution | Mean | SD | Median | | |
| 16 bits | 2.5 s | 22.6 s | 1.6 s | | |
| 14 bits | -1.6 s | 21.6 s | -3.7 s | | |
| 12 bits | -0.2 s | 22.9 s | -1.3 s | | |
| 10 bits | 0.5 s | 23.3 s | -3.5 s | | |
| 8 bits | 0.3 s | 24.7 s | -1.0 s | | |
| Linear mixed-effects model | | | | | |
| Digital resolution | *β* | *SE* | df | *t* ratio | *p* |
| 14 bits | -3.90 | 1.90 | 553 | -2.05 | .041* |
| 12 bits | -2.62 | 1.90 | 553 | -1.38 | .170 |
| 10 bits | -1.73 | 1.90 | 553 | -0.91 | .363 |
| 8 bits | -1.97 | 1.91 | 554 | -1.03 | .302 |
| Conditional $R^2$: 0.53; Marginal $R^2$: 0.003 | | | | | |
| Post-hoc contrasts | | | | | |
| Contrast | *p* | *d* | | | |
| 16 vs. 14 bits | .412 | 0.25 | | | |
| 16 vs. 12 bits | .604 | 0.17 | | | |
| 16 vs. 10 bits | .604 | 0.11 | | | |
| 16 vs. 8 bits | .604 | 0.13 | | | |
| 14 vs. 12 bits | .711 | -0.08 | | | |
| 14 vs. 10 bits | .604 | -0.14 | | | |
| 14 vs. 8 bits | .604 | -0.12 | | | |
| 12 vs. 10 bits | .799 | -0.06 | | | |
| 12 vs. 8 bits | .816 | -0.04 | | | |
| 10 vs. 8 bits | .899 | 0.02 | | | |

Table S16. Metrics on the impact of digital resolution. *$p < .05$

| *Sensitivity* | | | | | |
|---|---|---|---|---|---|
| Descriptives | | | | | |
| Number of channels | Mean | SD | Median | | |
| 4 | 80.2% | 26.9% | 100.0% | | |
| 3 | 77.3% | 26.1% | 85.7% | | |
| 2 | 72.2% | 31.1% | 83.3% | | |
| 1 | 71.2% | 22.8% | 75.0% | | |
| Linear mixed-effects model | | | | | |
| Number of channels | *β* | *SE* | df | *t* ratio | *p* |
| 3 | -2.94 | 1.68 | 432 | -1.75 | .081 |
| 2 | -8.10 | 1.68 | 432 | -4.83 | $1.93e^{-06}$* |
| 1 | -9.06 | 1.68 | 432 | -5.41 | $1.05e^{-07}$* |
| Conditional $R^2$: 0.72; Marginal $R^2$: 0.02 | | | | | |
| Post-hoc contrasts | | | | | |
| Contrast | *p* | *d* | | | |
| 4 vs. 3 channels | .097 | 0.21 | | | |
| 4 vs. 2 channels | < .0001* | 0.57 | | | |
| 4 vs. 1 channels | < .0001* | 0.64 | | | |
| 3 vs. 2 channels | .003* | 0.36 | | | |
| 3 vs. 1 channels | .001* | 0.43 | | | |
| 2 vs. 1 channels | .561 | 0.07 | | | |
| *False detections per hour* | | | | | |
| Descriptives | | | | | |
| Number of channels | Mean | SD | Median | | |
| 4 | 6.0 FD/h | 2.6 FD/h | 5.8 FD/h | | |
| 3 | 5.7 FD/h | 2.6 FD/h | 5.6 FD/h | | |
| 2 | 5.6 FD/h | 2.5 FD/h | 5.6 FD/h | | |

| | | | | | |
|---|---|---|---|---|---|
| 1 | 5.5 FD/h | 1.7 FD/h | 5.4 FD/h | | |
| Linear mixed-effects model | | | | | |
| Number of channels | *β* | *SE* | df | *t* ratio | *p* |
| 3 | -0.35 | 0.17 | 432 | -2.01 | .045* |
| 2 | -0.41 | 0.17 | 432 | -2.33 | .020* |
| 1 | -0.49 | 0.17 | 432 | -2.83 | .005* |
| Conditional $R^2$: 0.61; Marginal $R^2$: 0.01 | | | | | |
| Post-hoc contrasts | | | | | |
| Contrast | *p* | *d* | | | |
| 4 vs. 3 channels | .090 | 0.24 | | | |
| 4 vs. 2 channels | .060 | 0.27 | | | |
| 4 vs. 1 channels | .029* | 0.33 | | | |
| 3 vs. 2 channels | .747 | 0.04 | | | |
| 3 vs. 1 channels | .619 | 0.10 | | | |
| 2 vs. 1 channels | .743 | 0.06 | | | |
| *Average detection delay* | | | | | |
| Descriptives | | | | | |
| Number of channels | Mean | SD | Median | | |
| 4 | -2.1 s | 17.9 s | -2.5 s | | |
| 3 | 0.4 s | 19.2 s | -2.8 s | | |
| 2 | 2.5 s | 22.6 s | 1.6 s | | |
| 1 | 2.2 s | 15.2 s | 0.5 s | | |
| Linear mixed-effects model | | | | | |
| Number of channels | *β* | *SE* | df | *t* ratio | *p* |
| 3 | 2.42 | 1.83 | 421 | 1.32 | .187 |
| 2 | 4.58 | 1.85 | 424 | 2.47 | .014* |
| 1 | 4.25 | 1.82 | 422 | 2.33 | .020* |
| Conditional $R^2$: 0.34; Marginal $R^2$: 0.01 | | | | | |

| Post-hoc contrasts | | |
|---|---|---|
| Contrast | *p* | *d* |
| 4 vs. 3 channels | .363 | -0.16 |
| 4 vs. 2 channels | .061 | -0.30 |
| 4 vs. 1 channels | .061 | -0.28 |
| 3 vs. 2 channels | .363 | -0.14 |
| 3 vs. 1 channels | .377 | -0.12 |
| 2 vs. 1 channels | .857 | 0.02 |

Table S17. Metrics on the impact of number of channels. *$p < .05$
